# Altered heart-brain coupling in awake patients with isolated REM sleep behaviour disorder

**DOI:** 10.1101/2025.11.18.25340408

**Authors:** Fosco Bernasconi, Julia van der Meer, Zehra Merchant, Jevita Potheegadoo, Marzia De Lucia, Claudio Bassetti, Carolin Schaefer, Olaf Blanke

**Author notes:** These authors have contributed equally (Carolin Schaefer, Olaf Blanke). Corresponding authors:* Dr. Fosco Bernasconi, École Polytechnique Fédérale de Lausanne Neuro-X Institute, Campus Biotech H4.02 Ch. des Mines 9, 1202 Geneva, Prof. Olaf Blanke, École Polytechnique Fédérale de Lausanne Neuro-X Institute, Campus Biotech H4.02 Ch. des Mines 9, 1202 Geneva.

## Abstract

Isolated REM Sleep Behaviour Disorder (iRBD) represents an early stage of α-synucleinopathy, often preceding Parkinson’s disease, dementia with Lewy bodies or Multiple System Atrophy. Growing evidence shows that iRBD patients are not only characterized by sleep disturbances but also dysautonomia. However, whether the brain–body coupling reflecting the interaction between neural and peripheral physiological activity is altered in iRBD remains unknown. Leveraging whole-night polysomnography data from thirty-six participants, we quantified heart–brain coupling via heartbeat evoked potentials (HEPs) across wakefulness, NREM, and REM sleep. HEPs were compared between individuals with isolated REM sleep behavior disorder (iRBD; n = 13) and healthy controls (HC; n = 23). In addition, heart rate variability (HRV) and other ECG-derived features were analyzed. During wakefulness, iRBD patients showed altered HEP compared to HC, between 230 and 445 ms after the R-peak. over frontal regions. The duration of RBD symptoms was positively associated with the magnitude of these HEP alterations. HEP alterations were specific to wakefulness, as no differences were observed during NREM or REM sleep. Additional analyses showed that HEP alterations in iRBD during wakefulness were not driven by ECG differences. We corroborate previous findings of altered heart rate variability (HRV) in iRBD patients during REM sleep. We demonstrate that brain-body coupling, as indexed by HEP, is altered during wakefulness in iRBD patients. These diurnal HEP may represent a novel quantitative biomarker of iRBD, and could, in the future, serve as a marker for the phenoconversion to α-synucleinopathy.

## Introduction

Rapid eye movement (REM) sleep behaviour disorder (RBD) is characterized by vocalizations, jerks, and complex motor behaviours during REM sleep, often associated with vivid, negatively toned, and aggressive dream content, accompanied by the loss of normal muscle atonia (Schenck & Mahowald, 2002). In the absence of other neurological symptoms, RBD is defined as isolated (iRBD) and recognized as one of the earliest clinical manifestations of α-synucleinopathies (Högl et al., 2018; Howell et al., 2023). The prevalence of iRBD is documented to be approximately 1%, predominantly affecting older adults, particularly individuals over 60 years of age, and occurs more frequently in men than in women (Haba-Rubio et al., 2018). iRBD is a strong prodromal marker for Parkinson’s disease (PD), dementia with Lewy bodies (DLB), or rarely multiple system atrophy (Schenck & Mahowald, 2002, Postuma et al 2019). More than 80% of individuals with iRBD eventually develop one of these neurodegenerative disorders within a decade or more (Postuma et al., 2019). RBD is also a frequent feature of PD (e.g., Gagnon et al., 2002) and a core symptom of DLB (McKeith et al., 2017). In these cases, it is categorized as secondary RBD. In PD, the presence of RBD has been associated with a more aggressive course of the disease, characterized by faster progression and more severe non-motor symptoms (Berg et al., 2021).

Although iRBD is primarily associated with sleep disturbances, growing evidence also links autonomic dysfunction (i.e., dysautonomia) with iRBD (Iranzo et al., 2009). Given that iRBD constitutes an early manifestation of α-synucleinopathies, the co-occurrence or early appearance of autonomic dysfunction likely reflects the underlying spread of pathology within central autonomic networks (Iranzo et al., 2009). The spectrum of autonomic dysfunction in iRBD is broad. Clinical interviews revealed symptoms related to gastrointestinal, erectile, and urinary dysfunctions in iRBD (Claassen et al., 2010; Postuma et al., 2006, 2009) and different cardiovascular abnormalities such as cardiac sympathetic denervation (Miyamoto et al., 2006), abnormal baroreflex sensitivity, and reduced heart rate variability (HRV) during REM sleep (Ferini-Strambi et al., 1996; Postuma et al., 2010; Sorensen et al., 2012, 2013). Findings on HRV in iRBD patients are heterogeneous, with some studies reporting alterations in low-frequency and/or high-frequency HRV spectra, indicative of cardiac sympathetic and parasympathetic dysfunction, respectively (e.g., Yang et al., 2021). Other studies did not observe any abnormality in the HRV (Memon et al., 2024; Zitser et al., 2019).

Growing evidence shows that the brain integrates different interoceptive (bodily) signals arising from the internal organs (for review see Engelen et al., 2023). Hence, the neural activity is modulated by cardiac (e.g., De Falco et al., 2024; Park et al., 2017; Salomon et al., 2016), gastric (e.g., Mayeli et al., 2021; Rebollo et al., 2018) and breathing signals (e.g., Kluger et al., 2023; Park et al., 2020). The ascending interoceptive signals, such as those from the heart, that give rise to a neural response are conveyed to the brain via cranial nerves (especially the vagal nerve) as well as spinal pathways (Berntson & Khalsa, 2021). These ascending interoceptive signals are processed in the brainstem and propagate via the thalamus to viscerosensory cortices such as the insula, somatosensory, and anterior cingulate cortex (Berntson & Khalsa, 2021; A. D. B. Craig, 2009; De Falco et al., 2024; Garfinkel & Critchley, 2016; Park & Blanke, 2019). Even though the same regions and pathways, especially the brainstem and basal ganglia, are altered in iRBD patients (e.g., Andersen et al., 2025; Berg et al., 2021; Borghammer et al., 2021; De Marzi et al., 2016; Ehrminger et al., 2016; Hanyu et al., 2012; Rahayel et al., 2015; Scherfler et al., 2011; Unger et al., 2010) it is currently unknown whether the neural interoceptive processing is altered in these individuals.

Several lines of evidence suggest that awake patients with PD are characterized by impairments in interoceptive processing, either in showing reduced awareness of their own heartbeats (Hazelton et al., 2023; Ricciardi et al., 2016; Santangelo et al., 2018) or an altered neural response to heartbeat signal (Candia-Rivera et al., 2024). However, it has not been investigated whether these neural alterations in cardiac interoceptive processing are already present in iRBD (a prodromal stage of PD). Many recent studies investigating the brain processing of heartbeat signal in healthy individuals, analysed the so-called heartbeat-evoked potentials (HEPs), derived from electroencephalographic (EEG) signals that are time-locked to the R-peak of the electrocardiogram (ECG). HEPs typically manifest as a negative deflection approximately 200 to 600 milliseconds after the R-peak and are most prominent over fronto-central regions (e.g., Montoya et al., 1993; Park et al., 2016, 2017; Park & Blanke, 2019; Pollatos & Schandry, 2004).

Leveraging whole-night polysomnography recordings conducted in thirty-six individuals, we investigated whether the brain processing of the heartbeat signal, indexed by HEPs, is altered in patients with iRBD vs. healthy control individuals (HC), and whether this differed between different periods of the sleep-wake cycle (NREM and REM sleep, and during wakefulness). Based on the prior evidence mentioned above, we hypothesized that the HEP amplitude would be altered in awake iRBD individuals, but not necessarily during sleep. In addition, we also expected a difference (independent of the group) in HEP amplitude when comparing wakefulness with sleep.

## Materials and Methods

### Participants

Thirty-six datasets were included in the present analysis, comprising recordings obtained at the outpatient clinic of the Sleep–Wake–Epilepsy Center (SWEC) at Inselspital, University Hospital Bern, Switzerland. Thirteen datasets were from patients diagnosed with isolated REM sleep behavior disorder (iRBD), and twenty-three were from healthy control participants. All iRBD diagnoses were established by a neurologist specialized in sleep medicine. Exclusion criteria included a history of major psychiatric disorders, cerebrovascular disease, or any condition known to affect mental status. None of the participants had used antipsychotic medication within 5 years before the polysomnographic recording. The study protocol was approved by the local cantonal ethics committees (Bern BASEC #2022-01379 and 2021-00619; Geneva: #2017-01852).

### Polysomnography recordings and visual analysis

Overnight video-polysomnography (PSG) recordings were acquired using the *REMLogic* system (Embla Systems) and included bilateral frontal, central, temporal, and occipital electroencephalography (EEG) electrodes referenced online to Cz, bilateral electrooculography (EOG), and submental as well as anterior tibialis electromyography (EMG). PSG recordings were obtained between 2012 and 2021 and scored according to the criteria of the American Academy of Sleep Medicine (AASM 3). The diagnosis of iRBD was established according to the *International Classification of Sleep Disorders*, Third Edition (ICSD-III). In cases with absent or infrequent (≤1%) REM sleep, a diagnosis of probable iRBD was supported by a detailed clinical interview conducted by a trained sleep specialist. REM sleep behaviors were classified as minor, major, complex, or scenic, and vocalizations were documented. Visual scoring of N2 and N3 sleep stages was performed blinded to clinical information. For exploratory classification, PSGs were categorized as normal, moderately abnormal, or severely abnormal based on predefined criteria involving spindle density, slow-wave amplitude, and the persistence of wake-like EEG rhythms during NREM sleep.

### Automated sleep staging

To improve reproducibility and overcome inter-rater variability (Cesari et al., 2022), PSGs were automatically analysed to identify sleep stages using ‘Yet Another Spindle Algorithm’ (YASA) algorithm (Vallat & Walker, 2021). Sleep recordings were divided into 30-second epochs, and the previously validated algorithm with integral artifact rejection was used to automatically classify sleep stages based on scalp EEG (C4). The 30-s EEG epoch was classified as either wakefulness, N1 sleep, N2 sleep, N3 sleep, or REM sleep.

### Preprocessing

PSG recordings underwent a thorough artifact removal procedure; data were recorded with a sampling frequency of 200 Hz. First, the EEG continuous data were bandpass filtered between 0.5 Hz and 40 Hz (Butterworth of 5^th^ order) and then referenced to the two Mastoids. We epoched the continuous polysomnography data using the ECG R-peak as a time-locking event. Epochs included a window of -200ms and + 600ms centred on the R-peak. R-peaks were detected by applying MODWT with a sym4 wavelet to ECG signals. The squared reconstructed signal was analysed using MATLAB’s *findpeaks* function to identify R-peaks. Detected R-peaks were then inspected manually to exclude spurious events. Aberrant cardiac cycles with IBI < 0.4 or IBI > 1.5 (corresponding, respectively to a cardiac rate >120 bpm and <40 bpm) were excluded from all further computations. Epochs in which the EEG signal amplitudes were higher than ± 75 μV were rejected. Finally, epochs were visually inspected.

### Statistical analyses

#### Demographic and clinical variables

Statistical differences between iRBD and HC regarding demographic variables were assessed using the unpaired t-test and Fisher’s exact test. An unpaired t-test was employed to compare ECG features. RBD duration was calculated as the number of years between the PSG recording date and the onset of RBD symptoms reported by the patients. Because the onset of symptoms is not always precise (due to the subjective report from the patient), we set the beginning date to July 1 of the year indicated by each patient. For two patients, the duration was negative because the arbitrary onset date of July 1 occurred after the date of the PSG recordings. For these two patients, the duration was therefore set to zero.

#### HEPs

We computed the HEPs for each EEG channel (F3, F3, C3, C4, O1, O2) and sleep stage (awake, NREM, and REM). To assess HEP amplitude alterations in iRBD patients, we computed linear mixed models for each channel, time point, and sleep stage independently. We used amplitude as the dependent variable and group (iRBD vs. HC) and sleep stage (awake, NREM, and REM) as fixed effects and added an interaction between the two terms. Age was included as a covariate of no interest, and a random intercept was added for each individual to account for inter-individual variability. We also added a random slope for the sleep stage. The same analyses were computed for the ECG channel. Correction for multiple comparisons for the length of the epoch (-200ms to 600ms) was obtained with False Discovery Rate (FDR) (Benjamini & Yekutieli, 2001). During wakefulness we recorded 2’402 ± 990 (mean ± SD) HEPs for the iRBD and 2’533 ± 1’763 for HC (p-value = 0.777; t(1,34) = 0.285). During the NREM sleep stage, we recorded 12’858 ± 2’523 epochs for the iRBD and 13’145 ± 4’635 for HC (p-value = 0.812, t(1,34) = 0.24). In the REM phase, we observed 2’768 ± 1’726 epochs and 3’677 ± 1’814 for HC (p-value = 0.163, t(1,24) = 1.442).

## Results

### Polysomnographic recordings

We analysed polysomnography recordings of patients with iRBD (iRBD: n = 13) and healthy controls (HC: n = 23). The demographic data did not show significant (X^2^ = 1.853; p-value = 0.173) differences in sex (iRBD: 2/11 (female/male); HC: 8/15 (female/male) between the two groups, but we observed a significant difference (p-value = 0.01) in age, with patients with iRBD being older (iRBD: 65.7 ± 7 years; HC: 57.3 ± 8.8 years).

### Heartbeat-evoked potentials (HEPs)

To investigate alterations in the neural response to the heartbeat signal, we computed HEPs separately for each EEG channel (F3, F3, C3, C4, O1, O2) (Figure 1), sleep-wake cycle (NREM, REM, wakefulness), in each individual’s data set. As expected, visual inspection of the HEPs in each sleep stage showed a predominant response over fronto-central electrodes (Figures 1-2), and HEPs’ typical characteristics with a negative deflection in the EEG signal occurring approximately 200 to 400 milliseconds after the R-peak as defined based on the ECG recording (time-locking event).

**Figure 1.**
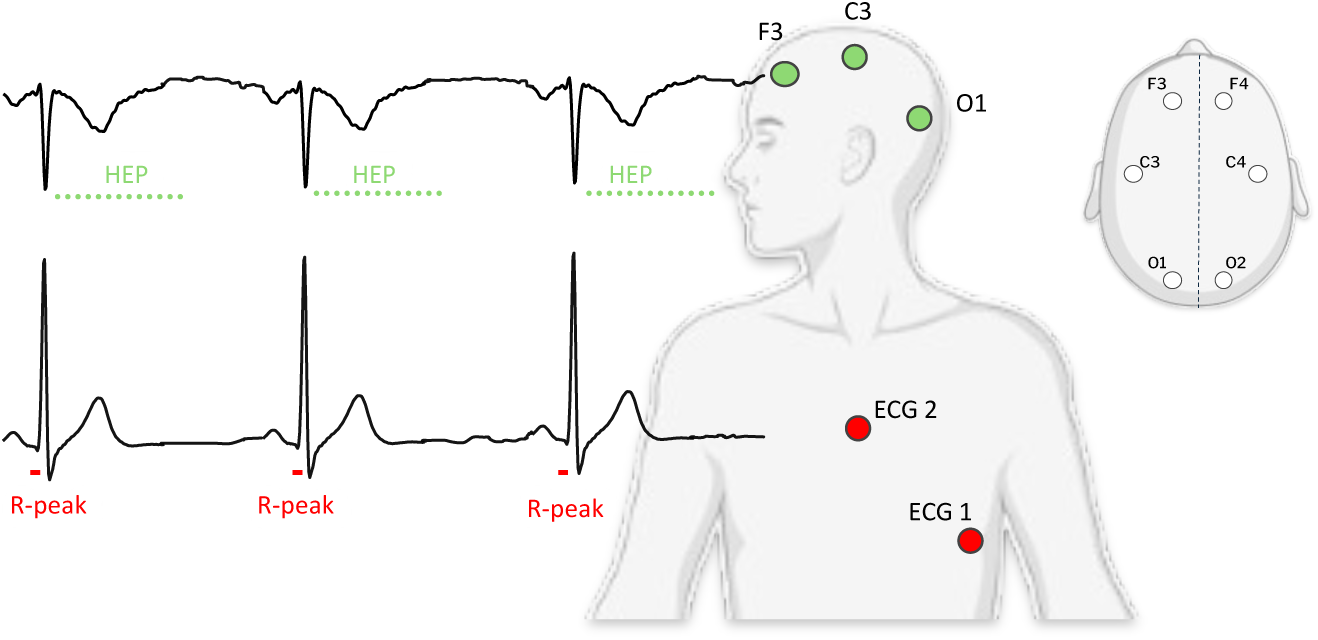
Polysomnography and HEPs. Illustration of the polysomnography setup, which includes the recording of the ECG (red dots), and six scalp EEG (green dots). HEPs are computed by time-locking the EEG signal to the R-peak of the ECG.

**Figure 2.**
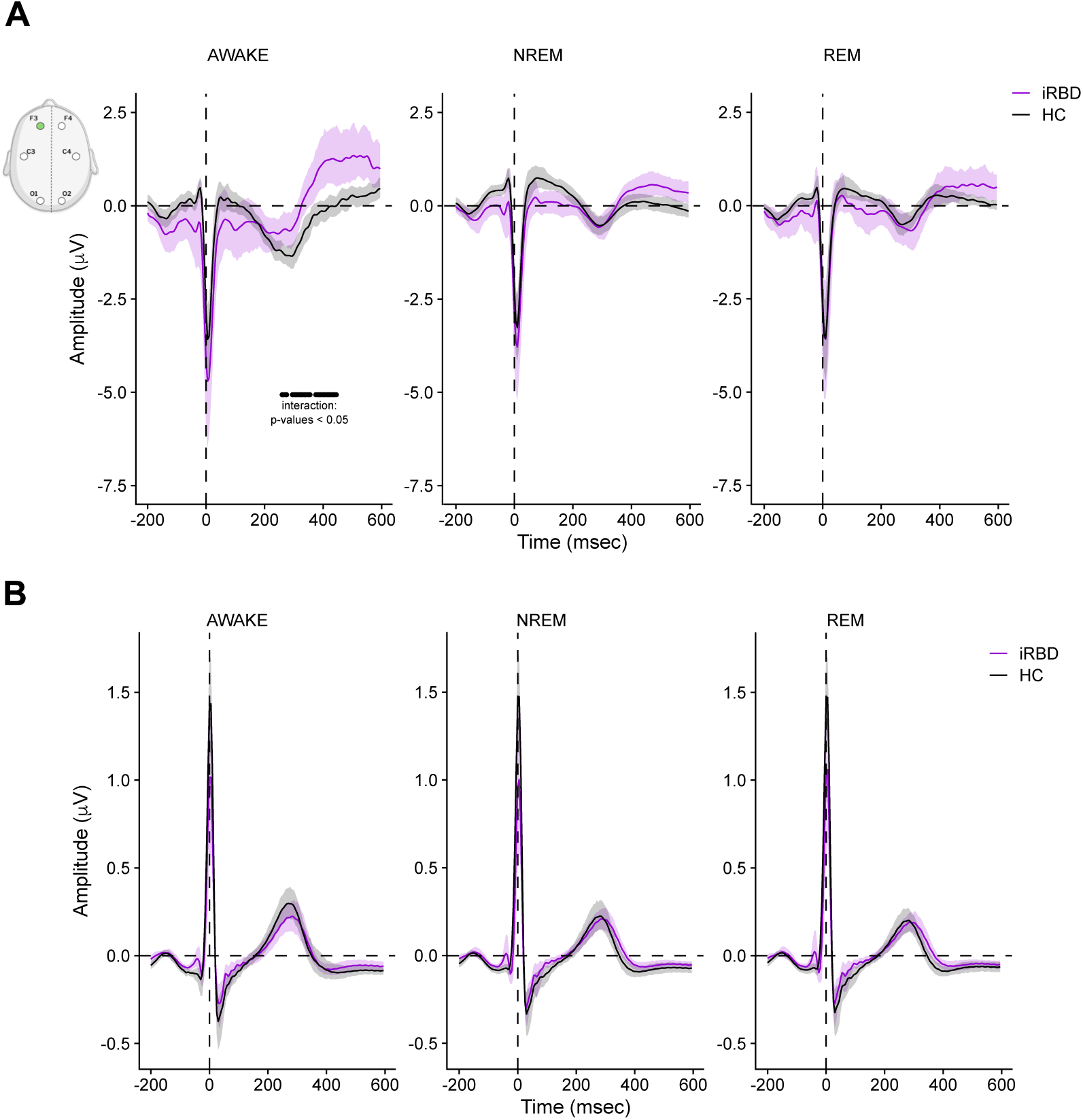
HEPs are altered in iRBD patients during wakefulness. **A.** HEPs on electrode F3 (green circle on the EEG layout). During wakefulness (left panel) are altered in iRBD patients (purple) compared to controls (black), the black line represents the period of significant interaction (Group, sleep stage; p-values < 0.05 and FDR-corrected) obtained through linear mixed models. The continuous lines represent the groups’ means, and the shaded areas represent the 95% CI. **B.** ECG amplitude. We did not observe any significant interaction (Group, sleep stage; p-values > 0.05; FDR-corrected).

To assess whether HEP amplitude differs between iRBD patients and HC, and whether such amplitude difference varies across sleep-wake stages, we fitted linear mixed-effects models for each channel and time point independently. HEP amplitude served as the dependent variable, with Group (iRBD vs. HC) and sleep–wake stage (NREM, REM, wakefulness) included as fixed effects, along with their interaction term. A random intercept was specified for each individual to account for inter-individual variability, and a random slope was included to model variability across sleep stages. Age was included as a covariate of no interest.

### HEPs are altered in iRBD during wakefulness, but not during NREM and REM sleep

Our results showed a significant interaction (p-values < 0.05, FDR-corrected) between *Groups* (iRBD vs. HC) and *sleep-wake stages* (wakefulness, NREM, and REM), over the period§ 260ms - 445ms after the R-peak (Figure 2A). This interaction was specific to the frontal electrode, lateralized on the left side (electrode F3). Post-hoc comparisons, computed on the mean of the whole significant interaction period (260ms - 445ms), showed that during wakefulness, iRBD patients have altered HEP amplitudes compared to controls (Figure 1A-B). This difference was not observed during NREM (Figure 1A) and REM sleep (Figure 1A-B). Previous studies in healthy individuals investigated whether nightmares are associated with HEP amplitude changes, showing no amplitude differences when comparing individuals with nightmares with those without (Bogdány et al., 2022; but see Perogamvros et al., 2019). Our results in iRBD patients align with these findings, showing no difference in HEP amplitude during sleep.

To exclude that these HEP differences (indexed by the interaction over the period 260ms - 445ms) might be related to ECG amplitude differences, we also investigated whether the ECG signal is modulated between the two subgroups and sleep stages (same statistical model as for the EEG electrodes). This analysis showed no statistically significant interaction (p-value > 0.05; FDR-corrected) (Figure 2B), suggesting that the altered HEPs observed in iRBD patients (Figure 2A-B) are not caused by a peripheral cardiac difference.

### Duration of RBD symptoms associated with HEP amplitude alteration

We investigated whether the duration of the RBD symptoms is associated with the HEP amplitude alterations observed during wakefulness in iRBD patients. With this aim, we conducted a robust regression (which present the advantage of being less affected by outliers (Maechler et al., 2025))between the duration of the RBD symptoms (from the onset of the symptoms till the day of the PSG; as dependent variable) and the HEP amplitude (mean of the 260ms - 445ms significant cluster for electrode F3). The mean duration of the RBD symptoms was 2.504 years (SD ± 2.760). Our results show a significant (F = 4.68, p-value = 0.05) positive association, with longer RBD symptoms duration being associated with stronger HEP amplitude alterations.

### HEP amplitudes are reduced across wake-sleep stages

Based on previous evidence from healthy young individuals showing that HEP amplitude is reduced during sleep when compared to wakefulness (Lechinger et al., 2015), we examined whether this modulation is also present in our elderly cohort. Our results indicate that during sleep, HEP amplitude is significantly (p-value > 0.05; FDR-corrected) modulated by the sleep-wake cycle (main effect), over all fronto-central electrodes (F3, F4, C3, and C4).

This modulation occurs during several distinct periods (Table 1; Figure 2). Post-hoc comparisons (calculated on the average of all significant timepoints for each channel separately) showed that HEP amplitude is decreased when comparing wakefulness, NREM, and REM sleep (main effect of sleep-wake stage). No statistical difference was observed between NREM and REM. These results corroborate and extend previous findings to a population of elderly individuals, showing that HEP amplitude decreases with sleep (Lechinger et al., 2015). Furthermore, the difference in HEP amplitude occurring before the R-peak has been associated with anticipatory mechanisms (Pelentritou et al., 2025), and is compatible with the view that the cardiac cycle, and therefore HEPs, is a continuous sequence of events without a distinct beginning or end.

**Table 1.** Significant cluster showing a modulation of HEPs amplitude as a function of sleep. The table indicates the periods during which a main effect of sleep was observed, for each electrode separately.

|  | Cluster 1 - Time (ms) | Cluster 2 - Time (ms) | Cluster 3 - Time (ms) |
| --- | --- | --- | --- |
| C3 | -95 - 310 | 495 - 530 | 540 - 595 |
| C4 | -85 - 15 | 35 - 290 | 480 - 595 |
| F3 | -155 - 10 | 30 - 310 | 470 - 595 |
| F4 | -190 - 290 | 465 - 595 |  |

To rule out the possibility that these HEP differences are related to ECG differences, we also assessed whether the ECG signal amplitude is influenced by sleep-wake stages. The findings reveal no statistically significant effect (p-value > 0.05; FDR-corrected) (Figure 3B), indicating that the changes in HEPs observed across sleep-wake stages are not simply due to peripheral cardiac differences.

**Figure 3.**
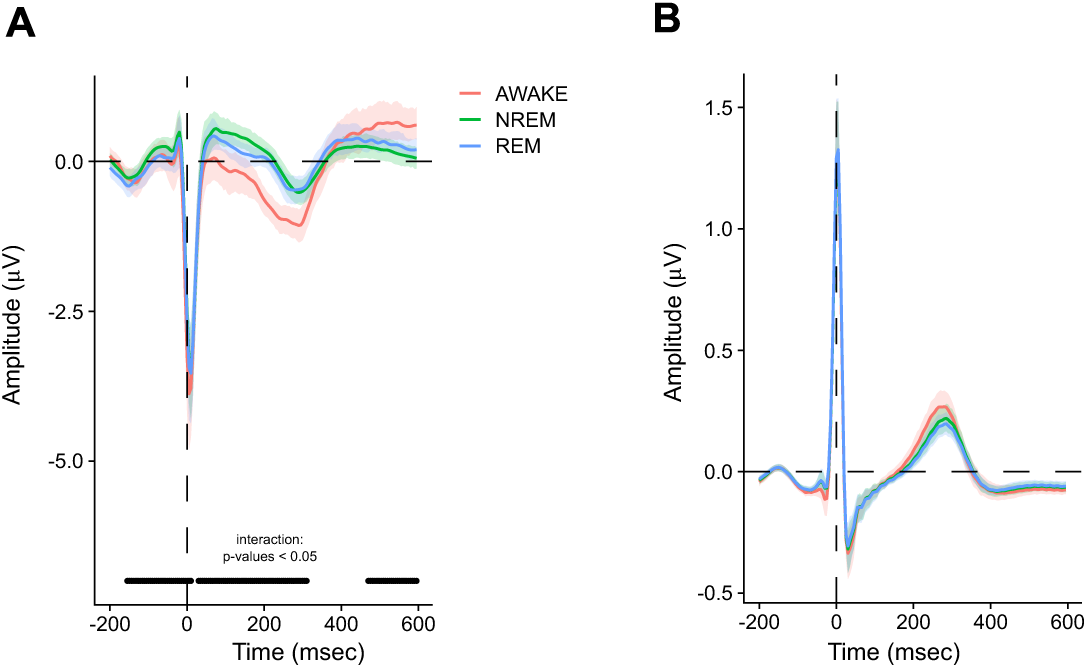
HEPs modulations across sleep-wake stages. **A.** HEPs on electrode F3 (part of the significant cluster), during wakefulness (red), NREM (green), and REM (blue). HEP amplitude is significantly reduced during NREM, and REM sleep in comparison to wakefulness. The horizontal black line represents the period of significant main effect of sleep (p-values < 0.05 and FDR-corrected) obtained through linear mixed models. The continuous lines represent the groups’ means, and the shaded areas represent the 95% CI. **B.** ECG amplitude. We did not observe any significant interaction (Group, sleep stage; p-values > 0.05; FDR-corrected).

### Heart rate variability (HRV) is reduced in iRBD during REM sleep

We also analysed several cardiac variables to corroborate previous results that are possibly modulated in iRBD, specifically during REM sleep (Ferini-Strambi et al., 1996; Gino et al., 2025; Lanfranchi et al., 2007; Postuma et al., 2009, 2010). During wakefulness, our results show the absence of statistically significant (all p-values > 0.05) differences between the two groups in heart rate (HR), inter-bit-interval (IBI), low and high power (absolute and relative) of HRV (see Table 2). Similar results were observed for the NREM sleep stage, for which we did not observe any significant difference (all p-values > 0.05) in cardiac variables. However, during the REM sleep stage, our results show a significant (p-values < 0.05) difference in high and low HRV absolute power in patients with iRBD compared to HC, replicating previous findings (Ferini-Strambi et al., 1996; Postuma et al., 2010; Sorensen et al., 2012, 2013). The other cardiac variables did not show any difference (p-values > 0.05) (Table 2).

**Table 2.** Demographic and cardiac features are modulated in iRBD patients, only during REM sleep stage. HR: heart rate variability; IBI: inter-beat-interval; HP: ECG relative high power; LP: ECG relative low power; LF: ECG absolute low power; HF: ECG absolute high power. Values for each variable represent the mean, and SD is indicated between parentheses.

| AWAKE | iRBD | HC | p | NREM | iRBD | HC | p | REM | iRBD | HC | p |
| --- | --- | --- | --- | --- | --- | --- | --- | --- | --- | --- | --- |
| HR | 57.6 (8.1) | 56.8 (16.6) | 0.87 | HR | 57.4 (8.1) | 57.6 (12.7) | 0.95 | HR | 58.2 (9.5) | 57.6 (14.1) | 0.9 |
| IBI | 1.1 (0.2) | 1.2 (0.6) | 0.38 | IBI | 1.1 (0.1) | 1.1 (0.4) | 0.64 | IBI | 1.1 (0.2) | 1.2 (0.6) | 0.53 |
| HP | 0.3 (0.2) | 0.3 (0.3) | 0.58 | HP | 0.3 (0.1) | 0.4 (0.1) | 0.77 | HP | 0.3 (0.2) | 0.4 (0.2) | **0.03** |
| LP | 0.7 (0.2) | 0.7 (0.3) | 0.77 | LP | 0.4 (0.2) | 0.4 (0.2) | 0.85 | LP | 0.7 (0.2) | 0.5 (0.2) | **0.05** |
| LF | 8.3 (8.5) | 4.1 (6.1) | 0.09 | LF | 114.0 (330.2) | 328.5 (1130.8) | 0.51 | LF | 1.0 (1.1) | 45.0 (146.6) | 0.31 |
| HF | 20.8 (29.1) | 71.8 (231.7) | 0.44 | HF | 71.7 (144.0) | 375.6 (1296.2) | 0.41 | HF | 2.2 (2.2) | 117.1 (379.4) | 0.31 |

## Discussion

iRBD is recognized as one of the earliest manifestations of α-synucleinopathy, often preceding the onset of PD or DLB. While emerging evidence points to potential alterations of interoceptive signals in patients with iRBD, this area remains largely unexplored. In the present study, leveraging whole-night polysomnography recordings, we analyzed brain processing of the heartbeat signal, reflected by HEPs recordings in iRBD patients vs. HC to determine the presence of altered brain-body coupling in iRBD patients. First, we report that during wakefulness, iRBD patients have altered HEP amplitude, but not during NREM and REM sleep stages (Figure 2). Second, the duration of the RBD symptoms is positively associated with the HEP amplitude alteration observed during wakefulness. Third, the HEP amplitude is decreased during sleep (compared to wakefulness; Figure 3). Fourth, we observed alterations in HRV in iRBD patients specifically during the REM sleep stage (Table 2). Taken together, our findings show sleep-wake stage-specific alterations in markers for brain-body coupling in iRBD patients. Such altered brain–body coupling may reflect underlying neuropathology and highlight HEP as a promising quantitative biomarker for iRBD.

Several pathophysiological mechanisms associated with iRBD may account for the altered HEPs we observed. First, iRBD has been associated with structural and functional alterations in the brainstem and thalamus (Boeve et al., 2007; Chen et al., 2013; García-Gomar et al., 2022; McKeith et al., 2017; Unger et al., 2010). These two brain regions have been identified as key subcortical hubs receiving afferent neural interoceptive signals coming from the heart (A. D. Craig, 2002; Critchley & Harrison, 2013; Garfinkel & Critchley, 2016; Park & Blanke, 2019). Therefore, we might interpret the altered HEP amplitude, observed in iRBD patients, as the result of the overlap between the core iRBD pathophysiological and the interoceptive processing pathways. Second, iRBD is also characterized by cortical alterations, including the sensorimotor cortex and frontal and posterior cortical regions (e.g., De Marzi et al., 2016; Hanyu et al., 2012; Rahayel et al., 2015). Some of these regions, especially the frontal, have been associated with HEP (e.g., Park et al., 2016; Park & Blanke, 2019), and their alterations might explain the altered HEP amplitude we observed in iRBD patients. Third, according to the recent body-first hypothesis, α-synuclein pathology may originate in peripheral organs and propagate to subcortical and cortical regions via the vagus nerve (Andersen et al., 2025; Borghammer, 2018; Borghammer et al., 2021; Dong et al., 2024; Horsager et al., 2022). The vagus nerve conveys interoceptive signals, such as those coming from the heart, from the organs to the brainstem; therefore, vagus nerve alterations due to the accumulation of α-synuclein might underlie the present HEP alterations. Fourth, the HEP alterations observed in our data might not only come from alterations in the brainstem and interoceptive pathways, but they might also arise due to the cardiac sympathetic denervation that has been observed in virtually every patient with iRBD (Kashihara et al., 2010; Knudsen et al., 2018; Miyamoto et al., 2006). Contributions by several of the indicated systems likely concur to produce the observed HEP changes in iRBD. Moreover, we note that our results showing HEPs alterations during wakefulness are not related to changes in the ECG signal nor ECG features (such as HRV). This is important and shows that the HEPs changes are rather related to alterations in neural interoceptive pathways.

Our results show no group differences in HEP amplitude during NREM and REM sleep. This may be explained by an overall reduction in HEP amplitude during these stages (compared to wakefulness), which could mask between-group differences due to the limited signal strength during sleep. Sleep-related HEP amplitude attenuation is thought to reflect reduced cortical responsiveness to visceral inputs, driven by state-dependent changes in thalamocortical dynamics and cortical excitability (Lechinger et al., 2015; Massimini et al., 2005).

We observed that iRBD have altered HRV during REM sleep stage, replicating previous findings (Ferini-Strambi et al., 1996; Gino et al., 2025; Lanfranchi et al., 2007; Postuma et al., 2009, 2010). While the underlying mechanisms for those HRV alterations during REM are not fully elucidated, it has been proposed that they might arise from the early cardiac sympathetic denervation, which becomes functionally apparent during REM sleep periods of autonomic instability, during which fluctuations between parasympathetic and increased sympathetic activity have been described. In contrast, during wakefulness, when the autonomic demand is relatively low, those alterations are usually not observed. There might also be some compensatory mechanisms (of the sympathetic nervous system) that may temporarily preserve normal HRV patterns despite the underlying pathology (Lanfranchi et al., 2007; Memon et al., 2024; Sorensen et al., 2013; Zitser et al., 2019). These selective HRV alterations during REM sleep might also be an indicator of early neurodegeneration in brainstem regions that coordinate both REM atonia and autonomic outflow (Lanfranchi et al., 2007). In conclusion, our results show that altered brain-heart interactions, indexed by HEPs, are a new quantitative marker for iRBD and the duration of the RBD symptoms. These results shed light on the possible use of HEP as a quick, accessible, and cost-effective marker for RBD. HEP alteration might already be present at the moment of early vagal symptoms, even in the absence of clinical RBD, possibly being also a prodromal marker.

Our findings should be considered in the context of the following limitations. First, the results are promising and should be replicated in a larger sample of patients. Second, although our results show effects with a reduced number of electrodes (n = 6), high-density EEG recordings would allow a more precise localisation of the HEP, as well as the possibility to further remove possible artefacts which might be present in the signals (e.g., pulse artifact). Third, longitudinal data are needed to corroborate the predictive role of HEP for the conversion to PD or DLB.

## Data Availability

Data will be made available via Gitlab after publication acceptance.

https://gitlab.epfl.ch/

## Acknowledgments

This research was supported by the Swiss National Science Foundation [grant n. 221182], Parkinson’s Schweiz Foundation, and the Leenaards Foundation to F.B.

## Data availability

Data will be made available via Gitlab after publication acceptance.

## Code availability

Code will be made available via Gitlab after publication acceptance.

## Authors contribution

Authors’ contributions according to the CRediT taxonomy (see http://credit.niso.org/ for more information).

Conceptualization: F.B. Validation: F.B.

Formal analysis: F.B., Z.M.

Investigation: J.VdM, C.S,

Ressources : F.B., C.S, C.B, O.B, J.P

Data curation: F.B., J.VdM

Writing – Original Draft: F.B. and O.B. Writing – Review & Editing: all authors

Visualization: F.B.

Project administration: F.B., O.B., C.S., J.VdM, J.P

Funding acquisition: F.B.; O.B

## Conflict of interest statement

The authors declare no conflict of interest.

## Notes

### Competing Interest Statement

The authors have declared no competing interest.

### Funding Statement

This research was supported by the Swiss National Science Foundation [grant 221182], Parkinson's Schweiz Foundation, and the Leenaards Foundation to F.B.

### Author Declarations

The study protocol was approved by the Swiss cantonal ethics committees (Bern BASEC #2022-01379 and 2021-00619; Geneva #2017-01852).

## References

Andersen, K. B., Krishnamurthy, A., Just, M. K., Van Den Berge, N., Skjærbæk, C., Horsager, J., Knudsen, K., Vogel, J. W., Toledo, J. B., Attems, J., Polvikoski, T., Saito, Y., Murayama, S., & Borghammer, P. (2025). Sympathetic and parasympathetic subtypes of body-first Lewy body disease observed in postmortem tissue from prediagnostic individuals. Nature Neuroscience, 28(5), 925–936. 10.1038/s41593-025-01910-9

Benjamini, Y., & Yekutieli, D. (2001). The control of the false discovery rate in multiple testing under dependency. The Annals of Statistics, 29(4), 1165–1188. 10.1214/aos/1013699998

Berg, D., Borghammer, P., Fereshtehnejad, S.-M., Heinzel, S., Horsager, J., Schaeffer, E., & Postuma, R. B. (2021). Prodromal Parkinson disease subtypes—Key to understanding heterogeneity. Nature Reviews Neurology, 17(6), 349–361. 10.1038/s41582-021-00486-9

Berntson, G. G., & Khalsa, S. S. (2021). Neural Circuits of Interoception. Trends in Neurosciences, 44(1), 17–28. 10.1016/j.tins.2020.09.011

Boeve, B. F., Silber, M. H., Ferman, T. J., Kokmen, E., Smith, G. E., Ivnik, R. J., Parisi, J. E., Olson, E. J., & Petersen, R. C. (1998). REM sleep behavior disorder and degenerative dementia: An association likely reflecting Lewy body disease. Neurology, 51(2), 363–370. 10.1212/wnl.51.2.363

Boeve, B. F., Silber, M. H., Saper, C. B., Ferman, T. J., Dickson, D. W., Parisi, J. E., Benarroch, E. E., Ahlskog, J. E., Smith, G. E., Caselli, R. C., Tippman-Peikert, M., Olson, E. J., Lin, S.-C., Young, T., Wszolek, Z., Schenck, C. H., Mahowald, M. W., Castillo, P. R., Del Tredici, K., & Braak, H. (2007). Pathophysiology of REM sleep behaviour disorder and relevance to neurodegenerative disease. Brain: A Journal of Neurology, 130(Pt 11), 2770–2788. 10.1093/brain/awm056

Bogdány, T., Perakakis, P., Bódizs, R., & Simor, P. (2022). The heartbeat evoked potential is a questionable biomarker in nightmare disorder: A replication study. NeuroImage: Clinical, 33, 102933. 10.1016/j.nicl.2021.102933

Borghammer, P. (2018). How does parkinson’s disease begin? Perspectives on neuroanatomical pathways, prions, and histology. Movement Disorders: Official Journal of the Movement Disorder Society, 33(1), 48–57. 10.1002/mds.27138

Borghammer, P., Horsager, J., Andersen, K., Van Den Berge, N., Raunio, A., Murayama, S., Parkkinen, L., & Myllykangas, L. (2021). Neuropathological evidence of body-first vs. Brain-first Lewy body disease. Neurobiology of Disease, 161, 105557. 10.1016/j.nbd.2021.105557

Candia-Rivera, D., Vidailhet, M., Chavez, M., & De Vico Fallani, F. (2024). A framework for quantifying the coupling between brain connectivity and heartbeat dynamics: Insights into the disrupted network physiology in Parkinson’s disease. Human Brain Mapping, 45(5), e26668. 10.1002/hbm.26668

Cesari, M., Heidbreder, A., St Louis, E. K., Sixel-Döring, F., Bliwise, D. L., Baldelli, L., Bes, F., Fantini, M. L., Iranzo, A., Knudsen-Heier, S., Mayer, G., McCarter, S., Nepozitek, J., Pavlova, M., Provini, F., Santamaria, J., Sunwoo, J.-S., Videnovic, A., Högl, B., … Stefani, A. (2022). Video-polysomnography procedures for diagnosis of rapid eye movement sleep behavior disorder (RBD) and the identification of its prodromal stages: Guidelines from the International RBD Study Group. Sleep, 45(3), zsab257. 10.1093/sleep/zsab257

Chen, M. C., Yu, H., Huang, Z.-L., & Lu, J. (2013). Rapid eye movement sleep behavior disorder. Current Opinion in Neurobiology, 23(5), 793–798. 10.1016/j.conb.2013.02.019

Claassen, D. O., Josephs, K. A., Ahlskog, J. E., Silber, M. H., Tippmann-Peikert, M., & Boeve, B. F. (2010). REM sleep behavior disorder preceding other aspects of synucleinopathies by up to half a century. Neurology, 75(6), 494–499. 10.1212/WNL.0b013e3181ec7fac

Craig, A. D. (2002). How do you feel? Interoception: the sense of the physiological condition of the body. Nature Reviews Neuroscience, 3(8), 655–666. 10.1038/nrn894

Craig, A. D. B. (2009). How do you feel--now? The anterior insula and human awareness. Nature Reviews. Neuroscience, 10(1), 59–70. 10.1038/nrn2555

Critchley, H. D., & Harrison, N. A. (2013). Visceral Influences on Brain and Behavior. Neuron, 77(4), 624–638. 10.1016/j.neuron.2013.02.008

De Falco, E., Solcà, M., Bernasconi, F., Babo-Rebelo, M., Young, N., Sammartino, F., Tallon-Baudry, C., Navarro, V., Rezai, A. R., Krishna, V., & Blanke, O. (2024). Single neurons in the thalamus and subthalamic nucleus process cardiac and respiratory signals in humans. Proceedings of the National Academy of Sciences, 121(11), e2316365121. 10.1073/pnas.2316365121

De Marzi, R., Seppi, K., Högl, B., Müller, C., Scherfler, C., Stefani, A., Iranzo, A., Tolosa, E., Santamarìa, J., Gizewski, E., Schocke, M., Skalla, E., Kremser, C., & Poewe, W. (2016). Loss of dorsolateral nigral hyperintensity on 3.0 tesla susceptibility-weighted imaging in idiopathic rapid eye movement sleep behavior disorder. Annals of Neurology, 79(6), 1026–1030. 10.1002/ana.24646

Dong, S., Shen, B., Jiang, X., Zhu, J., Zhang, H., Zhao, Y., Chen, Y., Li, D., Feng, Y., Chen, Y., Pan, Y., Han, F., Liu, B., & Zhang, L. (2024). Comparison of vagus nerve cross-sectional area between brain-first and body-first Parkinson’s disease. NPJ Parkinson’s Disease, 10, 231. 10.1038/s41531-024-00844-6

Ehrminger, M., Latimier, A., Pyatigorskaya, N., Garcia-Lorenzo, D., Leu-Semenescu, S., Vidailhet, M., Lehericy, S., & Arnulf, I. (2016). The coeruleus/subcoeruleus complex in idiopathic rapid eye movement sleep behaviour disorder. Brain: A Journal of Neurology, 139(Pt 4), 1180–1188. 10.1093/brain/aww006

Ej, O., Bf, B., & Mh, S. (2000). Rapid eye movement sleep behaviour disorder: Demographic, clinical and laboratory findings in 93 cases. Brain : A Journal of Neurology, 123 ( Pt 2). 10.1093/brain/123.2.331

Engelen, T., Solcà, M., & Tallon-Baudry, C. (2023). Interoceptive rhythms in the brain. Nature Neuroscience, 26(10), 1670–1684. 10.1038/s41593-023-01425-1

Ferini-Strambi, L., Oldani, A., Zucconi, M., & Smirne, S. (1996). Cardiac autonomic activity during wakefulness and sleep in REM sleep behavior disorder. Sleep, 19(5), 367–369. 10.1093/sleep/19.5.367

Gagnon, J. F., Bédard, M. A., Fantini, M. L., Petit, D., Panisset, M., Rompré, S., Carrier, J., & Montplaisir, J. (2002). REM sleep behavior disorder and REM sleep without atonia in Parkinson’s disease. Neurology, 59(4), 585–589. 10.1212/wnl.59.4.585

García-Gomar, M. G., Videnovic, A., Singh, K., Stauder, M., Lewis, L. D., Wald, L. L., Rosen, B. R., & Bianciardi, M. (2022). Disruption of Brainstem Structural Connectivity in REM Sleep Behavior Disorder Using 7 Tesla Magnetic Resonance Imaging. Movement Disorders: Official Journal of the Movement Disorder Society, 37(4), 847–853. 10.1002/mds.28895

Garfinkel, S. N., & Critchley, H. D. (2016). Threat and the Body: How the Heart Supports Fear Processing. Trends in Cognitive Sciences, 20(1), 34–46. 10.1016/j.tics.2015.10.005

Gino, C., Parati, M., Pincherle, A., Sattin, D., Dalla Vecchia, L. A., & De Maria, B. (2025). Advantage in sleep phase staging evaluating heart rate variability in patients with idiopathic rapid eye movement behaviour disorder: A systematic review with meta-analysis. European Journal of Preventive Cardiology, 32(Supplement_1), zwaf236.445. 10.1093/eurjpc/zwaf236.445

Haba-Rubio, J., Frauscher, B., Marques-Vidal, P., Toriel, J., Tobback, N., Andries, D., Preisig, M., Vollenweider, P., Postuma, R., & Heinzer, R. (2018). Prevalence and determinants of rapid eye movement sleep behavior disorder in the general population. Sleep, 41(2), zsx197. 10.1093/sleep/zsx197

Hanyu, H., Inoue, Y., Sakurai, H., Kanetaka, H., Nakamura, M., Miyamoto, T., Sasai, T., & Iwamoto, T. (2012). Voxel-based magnetic resonance imaging study of structural brain changes in patients with idiopathic REM sleep behavior disorder. Parkinsonism & Related Disorders, 18(2), 136–139. 10.1016/j.parkreldis.2011.08.023

Hazelton, J. L., Fittipaldi, S., Fraile-Vazquez, M., Sourty, M., Legaz, A., Hudson, A. L., Cordero, I. G., Salamone, P. C., Yoris, A., Ibañez, A., Piguet, O., & Kumfor, F. (2023). Thinking versus feeling: How interoception and cognition influence emotion recognition in behavioural-variant frontotemporal dementia, Alzheimer’s disease, and Parkinson’s disease. Cortex; a Journal Devoted to the Study of the Nervous System and Behavior, 163, 66–79. 10.1016/j.cortex.2023.02.009

Högl, B., Stefani, A., & Videnovic, A. (2018). Idiopathic REM sleep behaviour disorder and neurodegeneration—An update. Nature Reviews. Neurology, 14(1), 40–55. 10.1038/nrneurol.2017.157

Horsager, J., Knudsen, K., & Sommerauer, M. (2022). Clinical and imaging evidence of brain-first and body-first Parkinson’s disease. Neurobiology of Disease, 164, 105626. 10.1016/j.nbd.2022.105626

Howell, M., Avidan, A. Y., Foldvary-Schaefer, N., Malkani, R. G., During, E. H., Roland, J. P., McCarter, S. J., Zak, R. S., Carandang, G., Kazmi, U., & Ramar, K. (2023). Management of REM sleep behavior disorder: An American Academy of Sleep Medicine clinical practice guideline. Journal of Clinical Sleep Medicine: JCSM: Official Publication of the American Academy of Sleep Medicine, 19(4), 759–768. 10.5664/jcsm.10424

Iranzo, A., Molinuevo, J. L., Santamaría, J., Serradell, M., Martí, M. J., Valldeoriola, F., & Tolosa, E. (2006). Rapid-eye-movement sleep behaviour disorder as an early marker for a neurodegenerative disorder: A descriptive study. The Lancet Neurology, 5(7), Article 7. 10.1016/S1474-4422(06)70476-8

Iranzo, A., Santamaría, J., Rye, D. B., Valldeoriola, F., Martí, M. J., Muñoz, E., Vilaseca, I., & Tolosa, E. (2005). Characteristics of idiopathic REM sleep behavior disorder and that associated with MSA and PD. Neurology, 65(2), 247–252. 10.1212/01.wnl.0000168864.97813.e0

Iranzo, A., Santamaria, J., & Tolosa, E. (2009). The clinical and pathophysiological relevance of REM sleep behavior disorder in neurodegenerative diseases. Sleep Medicine Reviews, 13(6), 385–401. 10.1016/j.smrv.2008.11.003

Kashihara, K., Imamura, T., & Shinya, T. (2010). Cardiac 123I-MIBG uptake is reduced more markedly in patients with REM sleep behavior disorder than in those with early stage Parkinson’s disease. Parkinsonism & Related Disorders, 16(4), 252–255. 10.1016/j.parkreldis.2009.12.010

Kluger, D. S., Forster, C., Abbasi, O., Chalas, N., Villringer, A., & Gross, J. (2023). Modulatory dynamics of periodic and aperiodic activity in respiration-brain coupling. Nature Communications, 14(1), 4699. 10.1038/s41467-023-40250-9

Knudsen, K., Fedorova, T. D., Hansen, A. K., Sommerauer, M., Otto, M., Svendsen, K. B., Nahimi, A., Stokholm, M. G., Pavese, N., Beier, C. P., Brooks, D. J., & Borghammer, P. (2018). In-vivo staging of pathology in REM sleep behaviour disorder: A multimodality imaging case-control study. The Lancet. Neurology, 17(7), 618–628. 10.1016/S1474-4422(18)30162-5

Lanfranchi, P. A., Fradette, L., Gagnon, J.-F., Colombo, R., & Montplaisir, J. (2007). Cardiac Autonomic Regulation During Sleep in Idiopathic REM Sleep Behavior Disorder. Sleep, 30(8), 1019–1025. 10.1093/sleep/30.8.1019

Lechinger, J., Heib, D. P. J., Gruber, W., Schabus, M., & Klimesch, W. (2015). Heartbeat-related EEG amplitude and phase modulations from wakefulness to deep sleep: Interactions with sleep spindles and slow oscillations. Psychophysiology, 52(11), 1441–1450. 10.1111/psyp.12508

Maechler, M., Rousseeuw, P., Croux, C., Todorov, V., Ruckstuhl, A., Salibian-Barrera, M., Verbeke, T., Koller, M., Conceicao, E., & Anna di Palma, M. (2025). robustbase: Basic Robust Statistics. Http://Robustbase.r-Forge.r-Project.Org/.

Massimini, M., Ferrarelli, F., Huber, R., Esser, S. K., Singh, H., & Tononi, G. (2005). Breakdown of Cortical Effective Connectivity During Sleep. Science, 309(5744), 2228–2232. 10.1126/science.1117256

Mayeli, A., Zoubi, O. A., White, E. J., Chappelle, S., Kuplicki, R., Smith, R., Feinstein, J. S., Bodurka, J., Paulus, M. P., & Khalsa, S. S. (2021). Neural indicators of human gut feelings (p. 2021.02.11.430867). 10.1101/2021.02.11.430867

McKeith, I. G., Boeve, B. F., Dickson, D. W., Halliday, G., Taylor, J.-P., Weintraub, D., Aarsland, D., Galvin, J., Attems, J., Ballard, C. G., Bayston, A., Beach, T. G., Blanc, F., Bohnen, N., Bonanni, L., Bras, J., Brundin, P., Burn, D., Chen-Plotkin, A., … Kosaka, K. (2017). Diagnosis and management of dementia with Lewy bodies: Fourth consensus report of the DLB Consortium. Neurology, 89(1), 88–100. 10.1212/WNL.0000000000004058

Memon, A. A., George, E. B., Nazir, T., Sunkara, Y., Catiul, C., & Amara, A. W. (2024). Heart rate variability during sleep in synucleinopathies: A review. Frontiers in Neurology, 14, 1323454. 10.3389/fneur.2023.1323454

Miyamoto, T., Miyamoto, M., Inoue, Y., Usui, Y., Suzuki, K., & Hirata, K. (2006). Reduced cardiac 123I-MIBG scintigraphy in idiopathic REM sleep behavior disorder. Neurology, 67(12), 2236–2238. 10.1212/01.wnl.0000249313.25627.2e

Montoya, P., Schandry, R., & Müller, A. (1993). Heartbeat evoked potentials (HEP): Topography and influence of cardiac awareness and focus of attention. Electroencephalography and Clinical Neurophysiology, 88(3), 163–172. 10.1016/0168-5597(93)90001-6

Park, H.-D., Bernasconi, F., Bello-Ruiz, J., Pfeiffer, C., Salomon, R., & Blanke, O. (2016). Transient Modulations of Neural Responses to Heartbeats Covary with Bodily Self-Consciousness. Journal of Neuroscience, 36(32), 8453–8460. 10.1523/JNEUROSCI.0311-16.2016

Park, H.-D., Bernasconi, F., Salomon, R., Tallon-Baudry, C., Spinelli, L., Seeck, M., Schaller, K., & Blanke, O. (2017). Neural Sources and Underlying Mechanisms of Neural Responses to Heartbeats, and their Role in Bodily Self-consciousness: An Intracranial EEG Study. Cerebral Cortex (New York, N.Y.: 1991), 1–14. 10.1093/cercor/bhx136

Park, H.-D., & Blanke, O. (2019). Heartbeat-evoked cortical responses: Underlying mechanisms, functional roles, and methodological considerations. NeuroImage, 197, 502–511. 10.1016/j.neuroimage.2019.04.081

Pelentritou, A., Pfeiffer, C., Iten, M., Haenggi, M., Zubler, F., Schwartz, S., & De Lucia, M. (2025). Cardiac signals inform auditory regularity processing in the absence of consciousness. Proceedings of the National Academy of Sciences, 122(20), e2505454122. 10.1073/pnas.2505454122

Perogamvros, L., Park, H.-D., Bayer, L., Perrault, A. A., Blanke, O., & Schwartz, S. (2019). Increased heartbeat-evoked potential during REM sleep in nightmare disorder. NeuroImage. Clinical, 22, 101701. 10.1016/j.nicl.2019.101701

Plazzi, G., Corsini, R., Provini, F., Pierangeli, G., Martinelli, P., Montagna, P., Lugaresi, E., & Cortelli, P. (1997). REM sleep behavior disorders in multiple system atrophy. Neurology, 48(4), 1094–1097. 10.1212/wnl.48.4.1094

Pollatos, O., & Schandry, R. (2004). Accuracy of heartbeat perception is reflected in the amplitude of the heartbeat-evoked brain potential. Psychophysiology, 41(3), 476–482. 10.1111/1469-8986.2004.00170.x

Postuma, R. B., Gagnon, J. F., Vendette, M., & Montplaisir, J. Y. (2009). Markers of neurodegeneration in idiopathic rapid eye movement sleep behaviour disorder and Parkinson’s disease. Brain: A Journal of Neurology, 132(Pt 12), 3298–3307. 10.1093/brain/awp244

Postuma, R. B., Iranzo, A., Hu, M., Högl, B., Boeve, B. F., Manni, R., Oertel, W. H., Arnulf, I., Ferini-Strambi, L., Puligheddu, M., Antelmi, E., Cochen De Cock, V., Arnaldi, D., Mollenhauer, B., Videnovic, A., Sonka, K., Jung, K.-Y., Kunz, D., Dauvilliers, Y., … Pelletier, A. (2019). Risk and predictors of dementia and parkinsonism in idiopathic REM sleep behaviour disorder: A multicentre study. Brain: A Journal of Neurology, 142(3), 744–759. 10.1093/brain/awz030

Postuma, R. B., Lanfranchi, P. A., Blais, H., Gagnon, J.-F., & Montplaisir, J. Y. (2010). Cardiac autonomic dysfunction in idiopathic REM sleep behavior disorder. Movement Disorders: Official Journal of the Movement Disorder Society, 25(14), 2304–2310. 10.1002/mds.23347

Postuma, R. B., Lang, A. E., Massicotte-Marquez, J., & Montplaisir, J. (2006). Potential early markers of Parkinson disease in idiopathic REM sleep behavior disorder. Neurology, 66(6), 845–851. 10.1212/01.wnl.0000203648.80727.5b

Rahayel, S., Montplaisir, J., Monchi, O., Bedetti, C., Postuma, R. B., Brambati, S., Carrier, J., Joubert, S., Latreille, V., Jubault, T., & Gagnon, J.-F. (2015). Patterns of cortical thinning in idiopathic rapid eye movement sleep behavior disorder. Movement Disorders: Official Journal of the Movement Disorder Society, 30(5), 680–687. 10.1002/mds.25820

Rebollo, I., Devauchelle, A.-D., Béranger, B., & Tallon-Baudry, C. (2018). Stomach-brain synchrony reveals a novel, delayed-connectivity resting-state network in humans. eLife, 7, e33321. 10.7554/eLife.33321

Ricciardi, L., Ferrazzano, G., Demartini, B., Morgante, F., Erro, R., Ganos, C., Bhatia, K. P., Berardelli, A., & Edwards, M. (2016). Know thyself: Exploring interoceptive sensitivity in Parkinson’s disease. Journal of the Neurological Sciences, 364, 110–115. 10.1016/j.jns.2016.03.019

Salomon, R., Ronchi, R., Dönz, J., Bello-Ruiz, J., Herbelin, B., Martet, R., Faivre, N., Schaller, K., & Blanke, O. (2016). The Insula Mediates Access to Awareness of Visual Stimuli Presented Synchronously to the Heartbeat. The Journal of Neuroscience: The Official Journal of the Society for Neuroscience, 36(18), 5115–5127. 10.1523/JNEUROSCI.4262-15.2016

Santangelo, G., Vitale, C., Baiano, C., D’Iorio, A., Longo, K., Barone, P., Amboni, M., & Conson, M. (2018). Interoceptive processing deficit: A behavioral marker for subtyping Parkinson’s disease. Parkinsonism & Related Disorders, 53, 64–69. 10.1016/j.parkreldis.2018.05.001

Schenck, C. H., & Mahowald, M. W. (2002). REM sleep behavior disorder: Clinical, developmental, and neuroscience perspectives 16 years after its formal identification in SLEEP. Sleep, 25(2), 120–138. 10.1093/sleep/25.2.120

Scherfler, C., Frauscher, B., Schocke, M., Iranzo, A., Gschliesser, V., Seppi, K., Santamaria, J., Tolosa, E., Högl, B., Poewe, W., & SINBAR (Sleep Innsbruck Barcelona) Group. (2011). White and gray matter abnormalities in idiopathic rapid eye movement sleep behavior disorder: A diffusion-tensor imaging and voxel-based morphometry study. Annals of Neurology, 69(2), 400–407. 10.1002/ana.22245

Sorensen, G. L., Kempfner, J., Zoetmulder, M., Sorensen, H. B. D., & Jennum, P. (2012). Attenuated heart rate response in REM sleep behavior disorder and Parkinson’s disease. Movement Disorders: Official Journal of the Movement Disorder Society, 27(7), 888–894. 10.1002/mds.25012

Sorensen, G. L., Mehlsen, J., & Jennum, P. (2013). Reduced sympathetic activity in idiopathic rapid-eye-movement sleep behavior disorder and Parkinson’s disease. Autonomic Neuroscience, 179(1), 138–141. 10.1016/j.autneu.2013.08.067

Unger, M. M., Belke, M., Menzler, K., Heverhagen, J. T., Keil, B., Stiasny-Kolster, K., Rosenow, F., Diederich, N. J., Mayer, G., Möller, J. C., Oertel, W. H., & Knake, S. (2010). Diffusion tensor imaging in idiopathic REM sleep behavior disorder reveals microstructural changes in the brainstem, substantia nigra, olfactory region, and other brain regions. Sleep, 33(6), 767–773. 10.1093/sleep/33.6.767

Vallat, R., & Walker, M. P. (2021). An open-source, high-performance tool for automated sleep staging. eLife, 10, e70092. 10.7554/eLife.70092

Yang, J. H., Choi, S. H., Lee, M. H., Oh, S. M., Choi, J.-W., Park, J. E., Park, K. S., & Lee, Y. J. (2021). Association of heart rate variability with REM sleep without atonia in idiopathic REM sleep behavior disorder. Journal of Clinical Sleep Medicine : JCSM : Official Publication of the American Academy of Sleep Medicine, 17(3), 461–469. 10.5664/jcsm.8934

Zitser, J., During, E. H., Chiaro, G., & Miglis, M. G. (2019). Autonomic impairment as a potential biomarker in idiopathic REM-sleep-behavior disorder. Autonomic Neuroscience, 220, 102553. 10.1016/j.autneu.2019.05.005

